# Alcohol-related liver disease disrupts bile acid synthesis and is associated with compensatory gut microbiota changes

**DOI:** 10.1101/2025.07.23.25332046

**Authors:** Marisa Isabell Keller, Andressa de Zawadzki, Maja Thiele, Tommi Suvitaival, Karolina Sulek, Michael Kuhn, Christian Schudoma, Daniel Podlesny, Suguru Nishijima, Anthony Noel Fullam, Chan Yeong Kim, Lili Niu, Asger Wretlind, Johanne Kragh Hansen, Mads Israelsen, Wasiu Akanni, Diënty HM Hazenbrink, Helene Baek Juel, Matthias Mann, Torben Hansen, Aleksander Krag, Peer Bork, Cristina Legido-Quigley, GALAXY & MicrobLiver consortia

## Abstract

**Background:** Alcohol overuse disrupts liver function and alters microbial gut communities, with alcohol-related liver disease (ALD) accounting for half of all liver-related deaths worldwide. Bile acids (BAs) regulate liver and gut function, but their metabolism becomes disrupted in ALD. While it is known that gut microbes transform primary to secondary BAs, which are subsequently reabsorbed via the enterohepatic circulation, BA metabolism during ALD progression remains poorly understood.

**Methods:** We investigated BA co-metabolism in a cross-sectional cohort of individuals with ALD (n=462) and healthy controls (n=148). We validated key findings in two independent ALD cohorts (n=34 and n=52). We integrated BA concentrations, measured by targeted mass spectrometry in feces and plasma, with liver proteomics, and gut microbiome profiles derived from metagenomic and metatranscriptomic sequencing of fecal samples.

**Results:** Advanced fibrosis was associated with decreased hepatic BA synthesis and impaired BA transport. Despite this, disease progression corresponded with increased levels of primary and secondary BAs in plasma and feces. The abundance of microbial secondary BA dehydroxylation and epimerization pathways in the gut microbiome changed with disease severity. Genes encoding early steps in the multi-step dehydroxylation pathway increased, whereas those involved in later steps were depleted, indicating a community-level microbial imbalance. In ALD, we identified *Eggerthella lenta* as a key mediator of BA dehydroxylation, while *Mediterraneibacter torques* and *Bacteroides thetaiotaomicron* facilitated most of the BA epimerization as a detoxification mechanism.

**Conclusion:** Fibrotic ALD is characterized by disrupted primary BA synthesis and transport, leading to BA accumulation in the gut and blood circulation. Altered microbial secondary BA metabolism reflects a compensatory mechanism that becomes impaired at advanced fibrosis stages. Our findings highlight the gut-liver axis as an important factor influencing ALD progression.

**Impact and Implications:**

- With the progression of alcohol-related liver disease (ALD), levels of bile acids (BA) in serum and feces increase, but BA production and transport are impaired in the liver.
- Secondary microbial BA metabolism, particularly epimerization and dehydroxylation, increases in ALD. However, a key enzyme, *baiN,* is depleted, illustrating a microbial community-level metabolic dysbiosis.
- The main contributing microbial species were, among others, *Mediterraneibacter torques*, *Bacteroides thetaiotaomicron*, and *Eggerthella lenta*, which could serve as potential targets for future microbial-targeted interventions.

**Lay summary:** Alcohol-related liver disease (ALD) from long-term alcohol overuse affects how the liver and gut interact, especially in handling bile acids (BAs), which are molecules produced by the liver and transformed by gut bacteria. Our study found that in people with ALD, the liver produces fewer BAs, but BAs accumulate in the gut and blood because their transport is impaired. We also observed that bacterial transformations of these BA change as the disease progresses, most likely due to an imbalance in the gut microbiome. For the first time, we identify specific bacterial species that strongly influence BA levels in ALD.

Figure 0:
Graphical Abstract

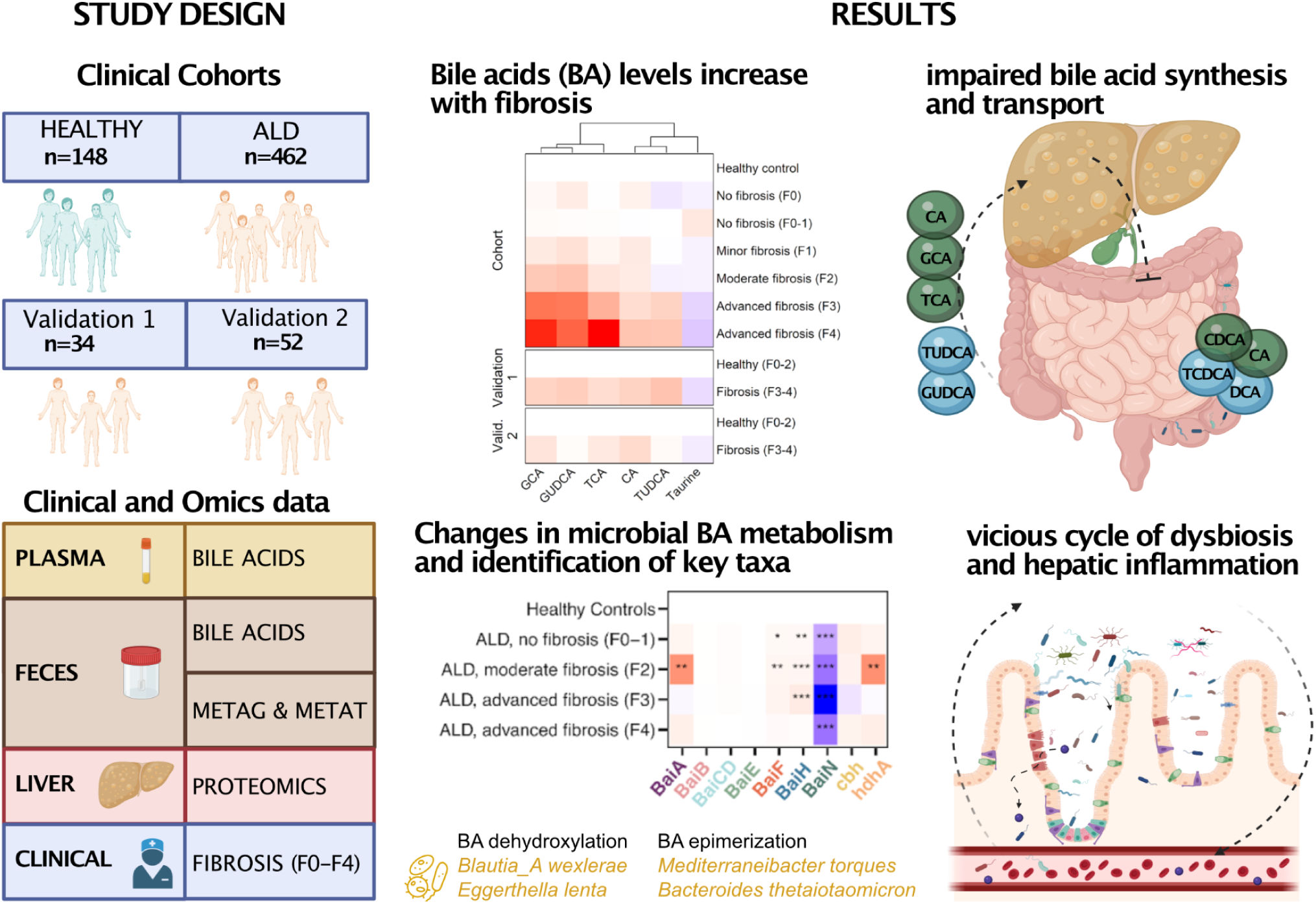

## Introduction

Alcohol-related liver disease (ALD) causes significant changes in the abundance of molecules produced or metabolised by the liver.^1^ Bile acid (BA) homeostasis protects the liver and other tissues from BA toxicity.^2^ This enterohepatic circulation balances BA synthesis and transport across different compartments and contributes to cellular homeostasis.^3^ Disruption of this equilibrium in liver disease results in abnormally high BA concentrations that are cytotoxic.^4^ Hydrophobic BAs act as detergents at elevated levels, causing misfolding of cytosolic proteins,^5^ disrupting cellular membranes,^6^ and inducing oxidative stress and DNA damage,^7^ thereby leading to hepatocyte injury, biliary duct damage, inflammation, cholestasis, and liver fibrosis.

The liver and the intestinal microbiome collaborate in digestion through biochemical pathways involving fat metabolism, vitamin production, carbohydrate digestion, and detoxification.^8^ Consequently, ALD-related metabolic changes have systemic effects and can be assessed via metabolite profiles in circulatory and excretory systems. BAs are synthesized by the liver from cholesterol and subsequently transported to the intestine, where they play a crucial role in fat digestion and vitamin absorption. Gut microbes convert primary BAs into secondary forms, which are then recycled by the liver.^9^ In the distal ileum, specific BA transporters reclaim up to 95% of the BAs, returning them to the liver via the portal vein, while approximately 5% are excreted in feces.^10^ Alcohol overuse in ALD is associated with alterations in the intestinal microbiome, which impact host metabolism.^11^ As primary BA production is regulated by receptors in the gut epithelium,^12^ gut microbes can inhibit BA production by modulating the expression of the 7-alpha-hydroxylase, the rate-limiting enzyme in BA synthesis.^13^ Moreover, microbial secondary BA metabolism, including (re)conjugation with taurine and glycine, dehydrogenation, epimerization, and dehydroxylation, alters BA toxicity and intestinal permeability.^14^

Although studies have measured BAs using liquid chromatography and mass spectrometry, they often failed to integrate host and microbial BA metabolism.^15^ Prior work linking 16S rRNA gene-based microbiome profiles to secondary BA metabolism capacity relied on correlations or species-specific *in vitro* studies, frequently limited to culturable bacteria.^16–18^ However, bacterial metabolism can be environment-dependent and differ between *in vivo* and *in vitro* study conditions.^19^ Thus, correlations between bacterial abundances and BAs may reflect indirect effects. While the presence of specific genes and transcripts provides insight into BA metabolism, a gap remains in understanding which bacteria actively drive these processes.

Here, we aim to disentangle the contribution of microbial secondary BA metabolism to disrupted BA homeostasis in ALD. We integrated liver proteomics, serum and fecal targeted metabolomics, and fecal metagenomic and metatranscriptomic analyses to obtain a holistic view. To identify potential microbial species as biomarkers and targets for microbial-based interventions, we mapped KEGG orthologs to metagenome-assembled genomes (MAGs) from the same cohort, linking functional changes to specific microbial species by considering gene presence and prevalence within species.

## Materials and methods

### Cohort description

#### Discovery Cohort

This study was conducted using plasma and fecal samples from the previously published GALAXY cross-sectional cohort of individuals with a history of harmful drinking (men > 36 g/day, women >24 g/day EtOH for at least one year) with an average of 19 years of excess alcohol use.^20–22^ This cohort focuses on asymptomatic ALD across the early disease stages, excluding individuals with diagnosed chronic liver disease or evident signs of advanced, decompensated liver disease. Together with matched healthy controls in the GALAXY-HC cohort (alcohol consumption units <7 units/week), the GALAXY-ALD cohort was used as the main discovery cohort. The healthy control group had normal glucose metabolism and liver function tests, no chronic or metabolic diseases, did not use antibiotics or any medications in the past six months, except occasional mild pain relievers. The Danish Data Protection Agency (13/8204, 16/3492) and the ethics committee for the Region of Southern Denmark (ethical ID S-20120071, S-20160021, S-20170087 and ID S-20160006G) approved these cohorts. Odense University Hospital recruited participants between 2013 and 2018, and obtained informed consent from all participants before inclusion. The study followed the ethical principles of the Declaration of Helsinki in all methods involving participants.

#### Validation Cohorts

Baseline data from two additional ALD cohorts are used in this study as validation cohort 1^23^ and validation cohort 2^24^. Validation cohort 1 is a randomized, single-center, placebo-controlled, double-blind study of ALD patients with fibrosis F1-4 registered under EudraCT number 20214-001856-51. Validation cohort 2 is a randomized, controlled study of ALD patients with compensated advanced chronic ALD (F3-4 and/or vibration-controlled transient elastography, VCTE, ≥10 kPa) registered in ClinicalTrial.gov ID NCT03863730. While both cohorts included participants in the main cohort, we here only included data from the non-overlapping participants.

#### Disease severity

We staged the ALD participants from liver histology (biopsies with 17G Menghini suction needle; Hepafix, Braun) based on Kleiner fibrosis score^25^ as F0 (no fibrosis), F1 (portal or periportal fibrosis), F2 (perisinusoidal fibrosis in combination with portal or periportal fibrosis), F3 (bridging fibrosis), and F4 (cirrhosis). For 97 participants from the GALAXY-ALD cohort, no liver histology was performed as their vibration-controlled transient elastography (VCTE) indicated no or minimal fibrosis (FibroScan<6.0 kPa). Consequently, those participants were grouped into the F0-F1 group in the analysis. The ALD and HC study populations differed in most liver health-related parameters, including liver stiffness measured by VCTE, Controlled attenuation parameter (CAP), and liver blood tests (Table 1). The samples from patients in the two validation cohorts showed similar ALD characteristics, but higher Kleiner scores, thus more advanced fibrosis (Supplementary Table 1).

**Table 1:**
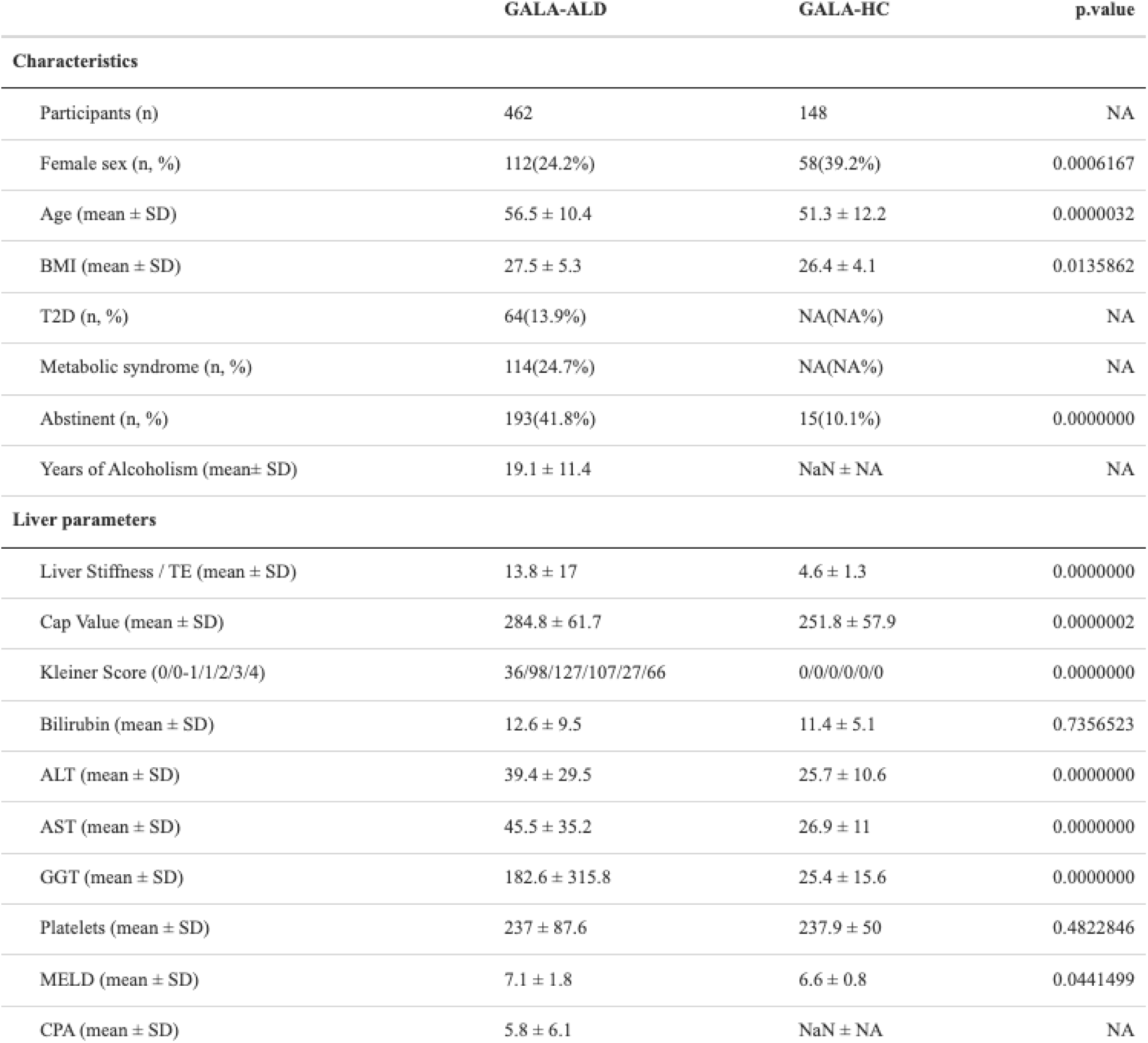
Characteristics of the GALAXY alcohol-related liver disease (ALD) and the GALAXY healthy controls (HC) cohort. T2D=Type 2 Diabetes, TE=transient elastography, Cap=Controlled attenuation parameter, ALT=alanine transaminase, AST=aspartate aminotransferase, GGT=gamma-glutamyl transferase, MELD=model of end-stage liver disease, CPA=C-reactive protein. The p-value reports the significance of the differences tested by the Kruskal-Wallis test for continuous variables and the chi-squared test for categorical variables.

|  | GALA-ALD | GALA-HC | p.value |
| --- | --- | --- | --- |
| **Characteristics** |  |  |  |
| Participants (n) | 462 | 148 | NA |
| Female sex (n, %) | 112(24.2%) | 58(39.2%) | 0.0006167 |
| Age (mean ± SD) | 56.5 ± 10.4 | 51.3 ± 12.2 | 0.0000032 |
| BMI (mean ± SD) | 27.5 ± 5.3 | 26.4 ± 4.1 | 0.0135862 |
| T2D (n, %) | 64(13.9%) | NA(NA%) | NA |
| Metabolic syndrome (n, %) | 114(24.7%) | NA(NA%) | NA |
| Abstinent (n, %) | 193(41.8%) | 15(10.1%) | 0.0000000 |
| Years of Alcoholism (mean ± SD) | 19.1 ± 11.4 | NaN ± NA | NA |
| **Liver parameters** |  |  |  |
| Liver Stiffness / TE (mean ± SD) | 13.8 ± 17 | 4.6 ± 1.3 | 0.0000000 |
| Cap Value (mean ± SD) | 284.8 ± 61.7 | 251.8 ± 57.9 | 0.0000002 |
| Kleiner Score (0/0-1/1/2/3/4) | 36/98/127/107/27/66 | 0/0/0/0/0/0 | 0.0000000 |
| Bilirubin (mean ± SD) | 12.6 ± 9.5 | 11.4 ± 5.1 | 0.7356523 |
| ALT (mean ± SD) | 39.4 ± 29.5 | 25.7 ± 10.6 | 0.0000000 |
| AST (mean ± SD) | 45.5 ± 35.2 | 26.9 ± 11 | 0.0000000 |
| GGT (mean ± SD) | 182.6 ± 315.8 | 25.4 ± 15.6 | 0.0000000 |
| Platelets (mean ± SD) | 237 ± 87.6 | 237.9 ± 50 | 0.4822846 |
| MELD (mean ± SD) | 7.1 ± 1.8 | 6.6 ± 0.8 | 0.0441499 |
| CPA (mean ± SD) | 5.8 ± 6.1 | NaN ± NA | NA |

#### Targeted Metabolomic Dataset

Quantitative BAs profiles were obtained from feces and plasma using a targeted method based on ultra-high-performance liquid chromatography coupled with a triple quadrupole mass spectrometer (UHPLC-MS/MS) as described previously ^26,27^. This method targets BAs as well as key metabolites relevant to metabolic diseases.

#### Liver Proteomics Dataset

The liver proteome was acquired as described previously^20^ and involved, in short, cryo-pulverizing liver biopsies, denaturing and digesting proteins with trypsin and LysC, acidifying to quench digestion, purifying peptides by solid-phase extraction, and drying them for concentration measurement. For LC–MS/MS analysis, 500 ng of purified peptides were injected and analyzed using data-independent acquisition on either an Evosep One LC system with an Orbitrap Exploris 480 mass spectrometer or an EASY-nLC 1200 system with a Q Exactive HF-X Orbitrap, with subsequent data analysis conducted using Spectronaut software and stringent filtering for statistical robustness.

#### Metagenomic and Metatranscriptomic of fecal samples

The metagenomic data were acquired as described previously^21^, and metatranscriptomics sequences were acquired in the same way. In short, the Qiagen All Prep Power Fecal DNA/RNA Kit with an additional phenol-chloroform step after cell lysis was used for DNA extraction, followed by library preparation using NEBNext Ultra II DNA library kit and dual index multiplex oligos while aiming for a 350-400 bp insert size. Sequencing of 2×150 bp paired-end reads was performed on an Illumina HiSeq 4000 platform. Using ngless^28^ (v1.1), we filtered out low-quality and host reads, keeping only bases with Phred >25 and reads >45 nt.

For microbial taxonomic profiles, the mOTUs^29^ v2.5 tool was used together with GTDB-tk^30^ (v2.11 r207) to acquire GTDB taxonomy for the ref-mOTUs via proGenomes2^31^ genomes. The meta-mOTUs were similarly mapped, and taxonomy was determined using a rule-based system and an 80% consistency threshold within clusters against the GTDB marker genes.

For microbial functional profiles, we followed two strategies. For in-depth gene-ontology information, the sequencing reads were aligned to the sub-catalog of the human gut microbiome within the global microbial gene catalog (GMGC)^32^ using BWA-MEM^33^ (v0.7.17). We used the eggNOG-mapper^34^ (v1.0.3) with the eggNOG database 5.0 to assign KEGG orthologies (KO) to each gene. Read counts for each KO were counted using gffquant (v2.9.1; https://github.com/cschu/gff_quantifier) while multiple mapping counts were split among the genes evenly. In parallel, the microbial functional pathway abundances were estimated by HumanN3 with a joint-index strategy^35,36^. We constructed metagenomic assembled genomes (MAG) as described before^37^ using and taxonomically classified them using GTDB-tk^30^ (v2.11 r207).

### Data analysis

Levels of metabolites in plasma were tested against the liver fibrosis stage using the R-package *limma* for an ANCOVA and post-hoc analysis, where metabolite level was explained by the fibrosis stage category. Metabolites, with any difference between the fibrosis stages, were tested further with pairwise comparisons between fibrosis stages. These metabolite patterns were validated in two external cohorts, where the agreement in effect sign was tested between the primary cohort and the two validation cohorts, each, with a binomial test. Finally, the estimated standardized effect sizes and metabolite levels concerning the fibrosis stage were visualized in a heatmap and boxplots, respectively. The same analysis strategy was repeated for metabolite levels in feces, as well as microbial taxa and microbial KEGG orthologues in feces.

Differential Abundance Analysis between two groups of patients was performed to investigate differences in the abundance of microbial pathways (abundance filter = 1e-7) in the fecal samples and proteins in the liver tissue (abundance filter = 1e-05). In the linear models and differential abundance analysis, the p-values were corrected for multiple testing using the Benjamini-Hochberg correction.^38^

## Results

### Levels of circulating and fecal bile acids increase during ALD progression

Targeted metabolomics of circulating metabolites revealed that ALD is associated with elevated plasma levels of the conjugated primary BAs taurocholic acid (TCA, Figure 1A), glycocholic acid (GCA, Figure 1B), and cholic acid (CA, Figure 1E), as well as the conjugated secondary BAs glycoursodeoxycholic acid (GUDCA, Figure 1C) and tauroursodeoxycholic acid (TUDCA, Figure 1D). Additionally, taurine, which is a substrate for BA conjugation, showed decreased plasma levels with advancing fibrosis (Figure 1F). These trends were confirmed in both validation cohorts (Figure 1F, binomial test for effect signs; p<0.05). Other measured metabolites did not show statistically significant changes (Supplementary Figure 1).

**Figure 1:**
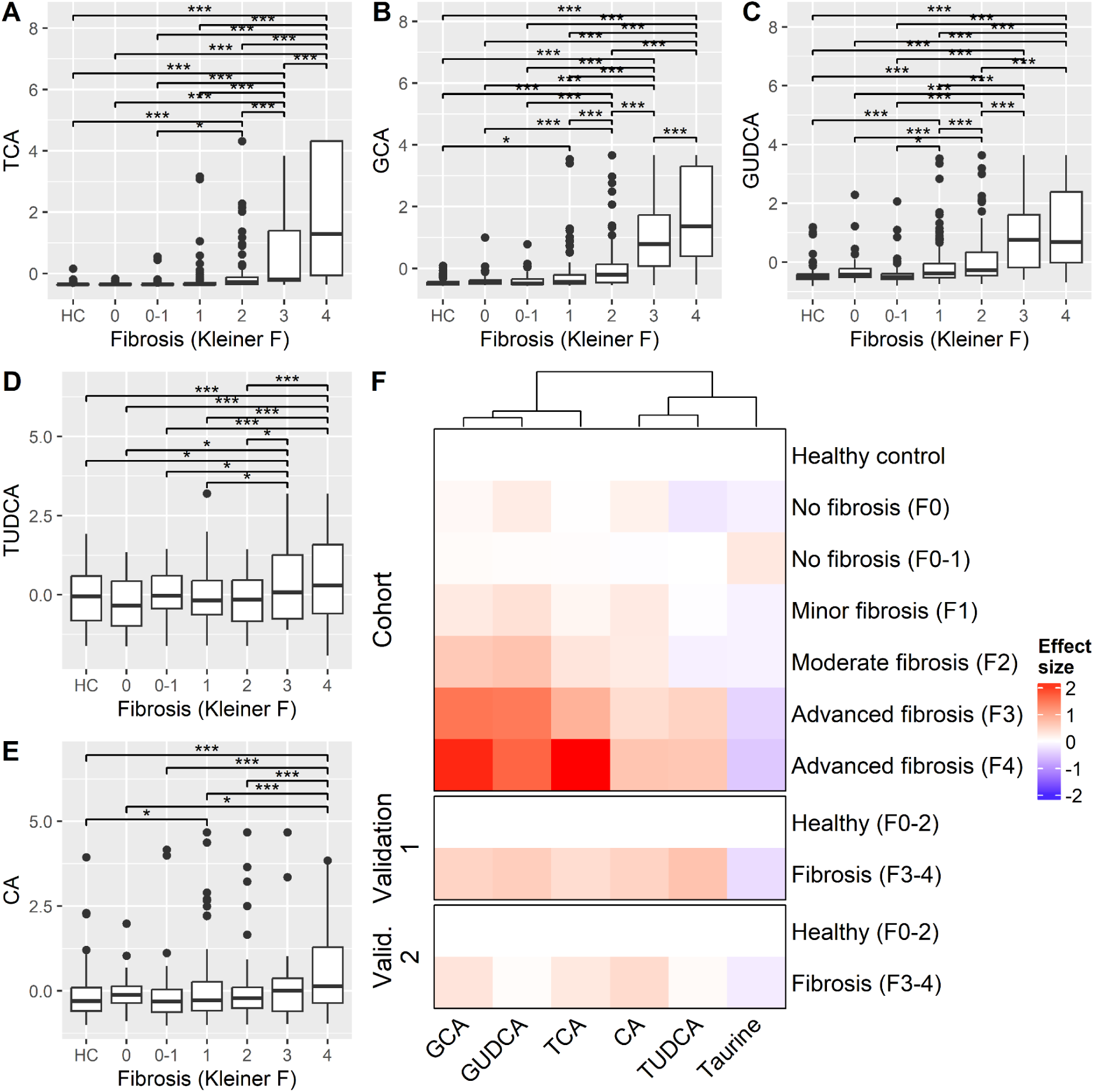
Serum metabolite levels according to fibrosis stage. **A-E.** Boxplot of standardized metabolite level (y-axis) grouped by fibrosis stage (x-axis) in the main cohort. Pairwise differences are highlighted as ***: multiple testing-corrected p<0.05; *: nominal p<0.05. **F.** Heatmap of the standardized effect size of metabolite level as compared to healthy control (main cohort) or ALD with no or minor fibrosis (validation cohorts). Differentially abundant metabolites are shown in columns (ordered by hierarchical clustering), fibrosis stages in rows (further grouped into panels according to cohort), and the respective effect size in color (red: increase; blue: decrease).

In fecal samples, the primary BA chenodeoxycholic acid (CDCA, Figure 2C) and its taurine-conjugated form taurochenodeoxycholic acid (TCDCA, Figure 2F) increased in abundance up to fibrosis stage three but declined at stage four. Similarly, cholic acid (CA, Figure 2A) and its secondary derivative deoxycholic acid (DCA, Figure 2E) showed increased levels with advancing fibrosis. While the trends for CDCA and DCA were consistent across the discovery and both validation cohorts, CA and TCDCA levels displayed heterogeneity across the validation cohorts. Additionally, azelaic acid decreased with fibrosis progression (Figure 2B). Other measured metabolites did not show statistically significant changes (Supplementary Figure 2).

**Figure 2:**
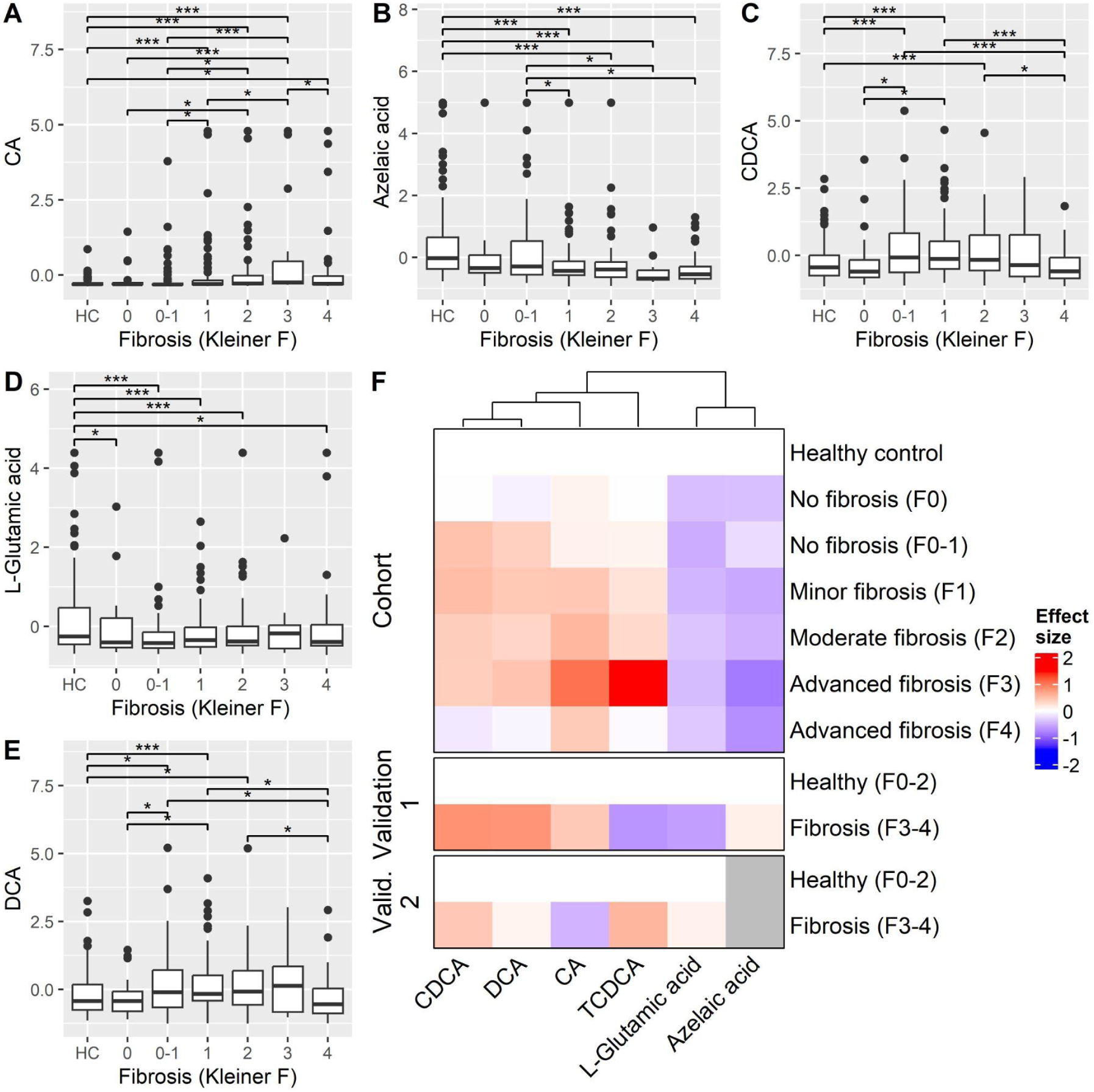
Fecal metabolite levels according to fibrosis stage. **A-E.** Boxplot of standardized metabolite level (y-axis) grouped by fibrosis stage (x-axis) in the main cohort. Pairwise differences are highlighted as ***: multiple testing-corrected p<0.05; *: nominal p<0.05. **F.** Heatmap of the standardized effect size of metabolite level as compared to healthy control (main cohort) or ALD with no or minor fibrosis (validation cohorts). Differentially abundant metabolites are shown in columns (ordered by hierarchical clustering), fibrosis stages in rows (further grouped into panels according to cohort), and the respective effect size in color (red: increase; blue: decrease; gray: not available).

### Dysregulation of human bile acid metabolism in ALD

The abnormal levels of primary and secondary conjugated BAs found in the plasma and feces of ALD patients indicate altered BA metabolism in the liver and impaired hepatic extraction. To investigate this, we analyzed 4,765 liver proteins measured by mass spectrometry and identified 1,146 proteins that were significantly different between low (F0-1) and high fibrosis (F2-4) groups (Supplementary Figure 3). Among these, four enzymes involved in primary BA biosynthesis were progressively downregulated with increasing liver fibrosis: CYP8B1, AKR1D1, BA-CoA-ligase (BAL, S27A5), and AKR1C4 (Figure 3). CYP8B1 (cytochrome P450 family 8 subfamily B member 1) encodes the enzyme 7-ɑ hydroxycholest-4-en-3-one 12-ɑ hydroxylase, a key protein responsible for synthesizing CA from cholesterol.^39^ AKR1D1 encodes aldo-keto reductase family 1 member D1, which is essential for the production of cholic acid.^40^

**Figure 3:**
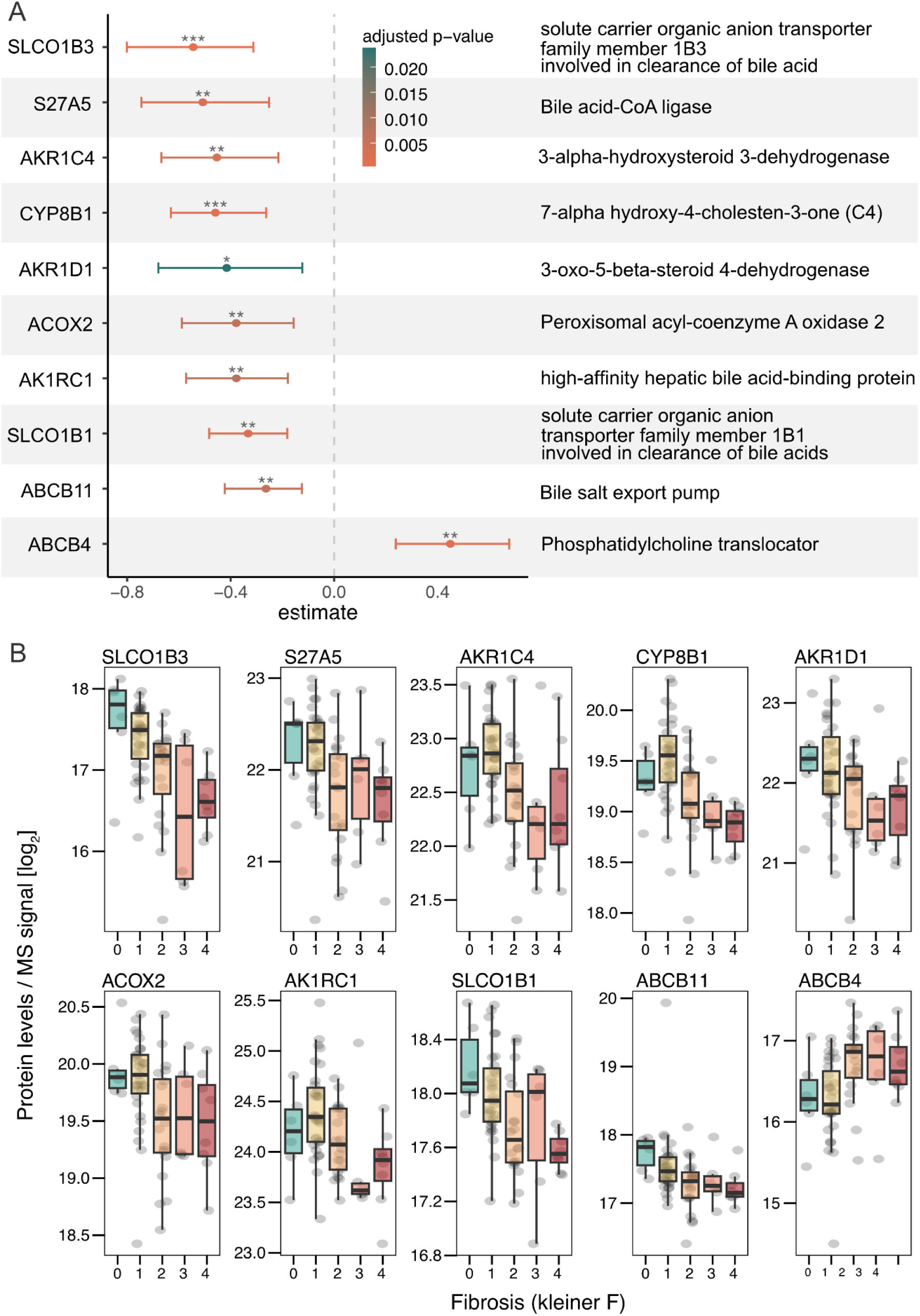
Liver protein changes their expression upon fibrosis. **A.** Liver proteins that significantly change between no or minor fibrosis (F0-1) and moderate or advanced fibrosis (F2-4) within the ALD cohort. The protein has been identified by differential abundance analysis using the Wilcoxon rank sum test and subsequent Benjamini-Hochberg correction for false discovery rate on the whole dataset with 4,765 liver proteins. Shown is the estimate with a confidence interval. Negative estimates correspond to a decrease in abundance upon fibrosis progression. **B.** Protein expression of the identified liver proteins from panel A is shown according to fibrosis stage.

Genes encoding solute carrier organic anion transporters of family 1B members (SLCO1B1 and SLCO1B3), which mediate hepatic uptake of endogenous compounds like BAs and bilirubin,^41^ were also downregulated with advancing fibrosis (Figure 3). Export mechanisms protecting hepatocytes from cytotoxic BA accumulation were impaired as well. Expression of the bile salt export pump (BSEP), encoded by ABCB11, decreased with fibrosis progression. BSEP is the main transporter of hydrophobic BAs across the canalicular membrane of hepatocytes.

Conversely, the phosphatidylcholine translocator (encoded by ABCB4) was significantly upregulated with increasing fibrosis. This protein facilitates the excretion of phospholipids from the liver into bile, where they form mixed BA-phospholipid micelles, protecting biliary epithelial cells from irritation caused by high concentrations of bile salts.^42^

### Microbial bile acid epimerization and dehydroxylation pathway abundances increase in ALD

Microbes in the human intestine play a major role in BA homeostasis, as they metabolise the bile acids to their secondary forms and are increasingly recognised as important modulators of liver diseases.^3^ Therefore, we investigated microbial functional profiles regarding their secondary bile acid metabolism. Among 549 microbial pathways detected in the discovery cohort, the BA epimerization and the 7-β and 7-ɑ dehydroxylation pathways were differentially enriched in ALD subjects compared to the healthy controls (Supplementary Figure 4A). Within the ALD cohort, comparing low fibrosis (F0-1) to high fibrosis (F2-4) groups, BA epimerization pathways increased in abundance with more advanced fibrosis (Supplementary Figure 4B).

We further analyzed the abundance of secondary bile acid–pathway gene orthologs in metagenomic data (metaG) and their expression in metatranscriptomic data (metaT). The multistep dehydroxylation of the primary BAs CA and CDCA to the secondary BAs DCA and lithocholic acid (LCA) by gut microbes is encoded by the BA-inducible (*bai*) genes.^17^ We found individual *bai* operon genes to be significantly altered in their abundance (Figure 4A-C). Genes encoding early oxidation steps, *baiA* (Figure 4G)*, baiH* (Figure 4H)*, baiF* (Figure 4F)*, and baiB* (Figure 4D), were enriched in ALD. Although statistical significance in the metatranscriptomic data was less robust than in metagenomic data, partly due to the lower sequencing depth and mapping rates, similar trends were generally observed. Transcription of the *baiB* and *baiA* genes was significantly higher in ALD patients than in healthy controls. In contrast, *baiN*, a gene encoding a late step in the oxidation pathway,^18^ showed the highest overall abundance in the microbiomes, but was strongly depleted in both gene and transcript levels with ALD and advanced fibrosis.

**Figure 4:**
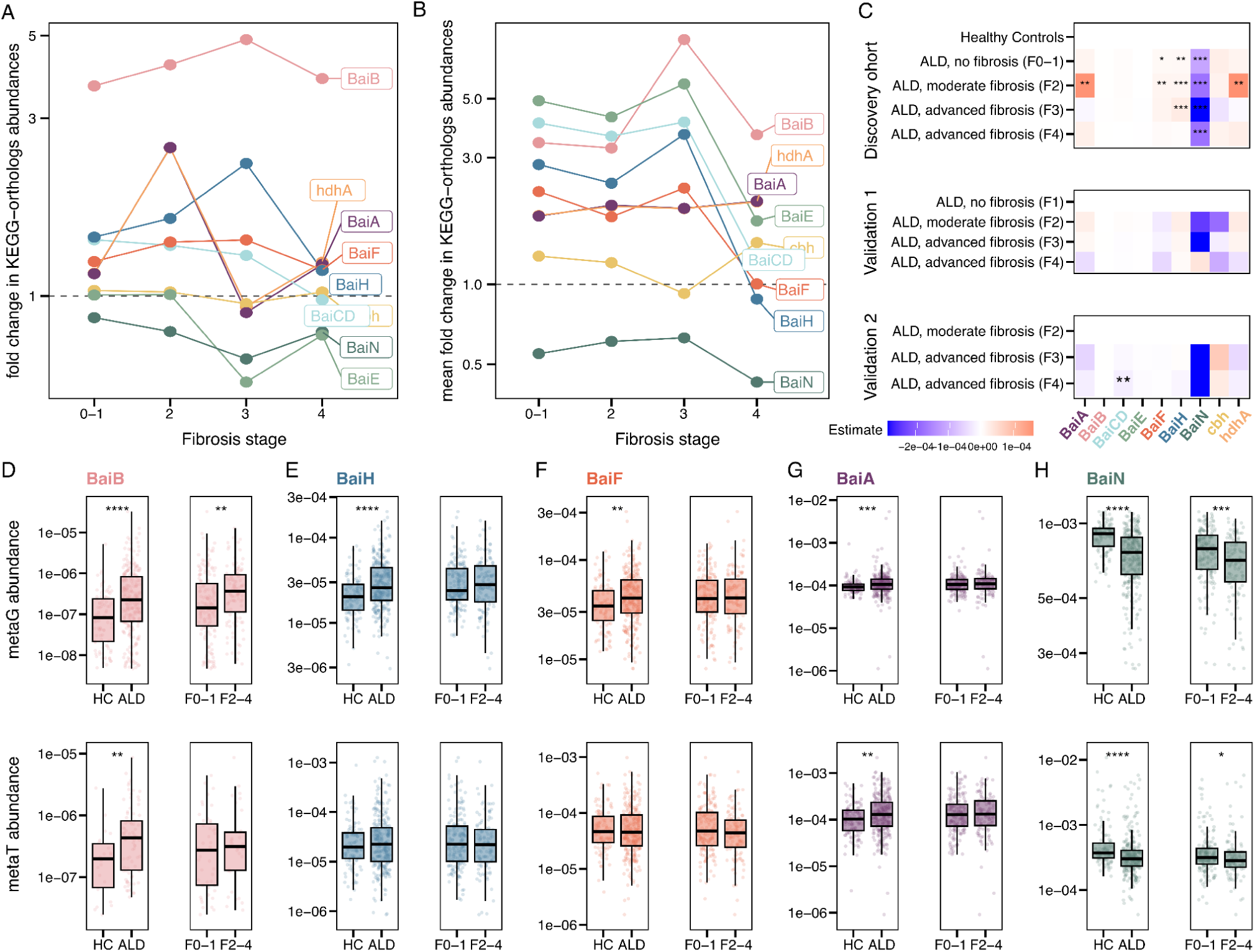
Microbial genes of the secondary BA metabolism change upon fibrosis stages. **A-B.** Abundance fold change of microbial secondary BA metabolism KEGG orthologs genes derived from metagenomic (metaG) sequencing (A) and gene expression derived from metatranscriptomic (metaT) sequencing (B) for each fibrosis stage in comparison to the healthy control samples. **C.** Heatmap of the linear model estimate of the abundance change for KEGG ortholog genes derived from metagenomic sequencing. Significance is reported after correcting for multiple testing using the Benjamini-Hochberg correction. *q<0.05, **q<0.01, ***q<0.001. **D-H.** Comparison of KEGG ortholog gene abundance (metaG) and gene expression (metaT) between healthy controls (CH) and samples from alcohol-related liver disease (ALD) and within the ALD group between no or minor fibrosis (F0-1) and moderate or advanced fibrosis (F2-4)

Enzymes encoded by 7-α-hydroxysteroid dehydrogenase (*hdhA*, also known as 7-α*-hsdh*) contribute to secondary BA diversity by promoting oxidation and epimerization, producing derivatives like ursocholic acid (UCA) and ursodeoxycholic acid (UDCA), including their taurine and glycine-conjugated forms.^14^ This pathway is favorable for both microbiota and host, as it detoxifies the BA pool by converting hydrophobic BAs into more hydrophilic forms.^43^ In the ALD cohort, the *hdhA* gene was notably upregulated, especially during early fibrosis stages (Figure 4C), but decreased in abundance again in advanced fibrosis, as observed in the validation cohorts. This U-shaped pattern from early to late fibrosis has also been detected for other molecules in previously and ongoing (unpublished) analyses of this cohort.^20^

### Identification of bacterial species most responsible for altered bile acid metabolism

To pinpoint the microbial species driving the metabolic changes during ALD progression, we mapped KEGG orthologous genes from our dataset onto metagenome-assembled genomes (MAGs) constructed from samples from the same discovery cohort of ALD and HC subjects. MAG assembly yielded 32,336 bins, of which 3,746 high-quality (>95% completeness, <5% contamination) and 15,914 medium-quality (>50% completeness, <10% contamination) bins were used for downstream analysis. This approach allowed us not only to assess species abundance fold changes but also to calculate the fraction of MAGs from each species carrying the genes of interest. Importantly, the fraction of gene-carrying MAGs should also show a high prevalence in our samples. We successfully mapped the *bai* genes with the highest overall abundance (*baiN*, K07007), the highest fold change (*baiB*, K15868), and the gene responsible for BA oxidation (*hdhA*, K00076).

In addition, we investigated the bile salt hydrolase gene (*bsh*; in KEGG choloylglycine hydrolase=*cbh*, K01442), as it encodes a key “gatekeeper” reaction in secondary BA metabolism. The deconjugation of glycine and taurine from primary BAs is a crucial initial step enabling subsequent BA transformations.^44^ The *bsh* gene is widely distributed across the bacterial tree of life^45^, reflected here by 753 species identified as carrying *bsh* within our study population (Figure 5A). While the overall abundance of *bsh* did not change (Figure 4C), we identified *Blautia_A wexlerae* and *Parabacteroides distasonis* as key species with a high fraction of gene-carrying MAGs and increased abundance during disease progression (Supplementary Figure 5).

**Figure 5:**
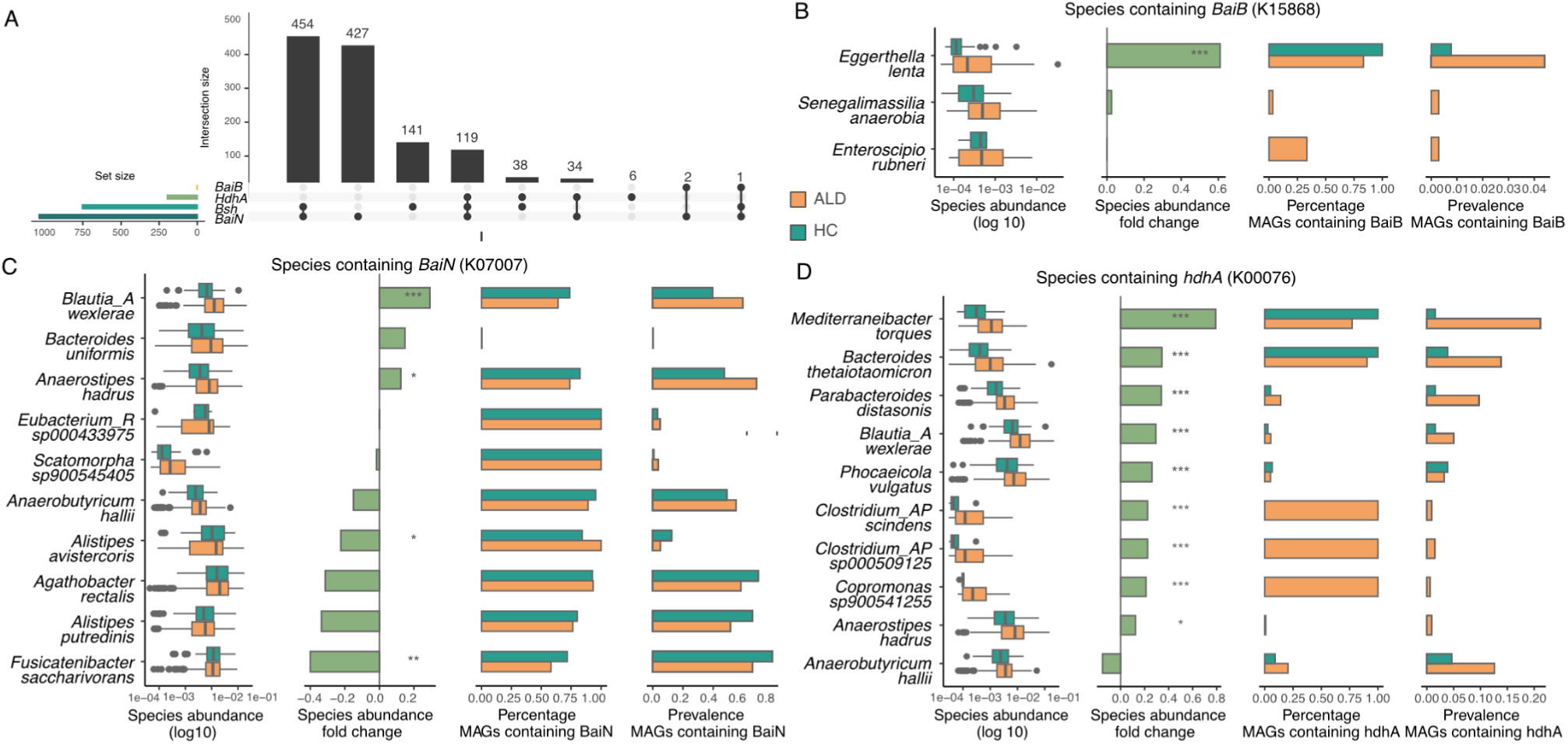
Importance of microbial taxa for secondary bile acid metabolism. **A.** Intersection of bacterial species containing the four genes of interest. **B-C.** Microbial taxa responsible for the secondary BA metabolism dehydroxylation pathway are facilitated by the *bai* operon. For *BaiB* (B), the gene was only found on MAGs from three different taxa. For the *BaiN (B)* gene, the ten species with the highest fold change abundance between ALD and HC are shown. **D.** Microbial taxa for secondary BA metabolism oxidation facilitated by *hdhA.* Importance is collectively estimated by the species abundance, species fold change between healthy and ALD samples, the gene-carrying frequency of MAGs, and the prevalence of gene-carrying MAGs in the cohort. Significance levels in the fold change are derived from a differential abundance analysis and corrected for multiple testing: *p<=0.05, **p<= 0.01, ***p<= 0.001.

### *Eggerthella lenta* facilitates the early arm of the BA dehydroxylation pathway, while the late pathway gene BaiN is prevalent across many taxa

The CoA ligase *baiB* gene, encoding the first enzyme of the oxidation arm of the dehydroxylation pathway, was originally identified in *Clostridium hylemonea* and *Clostridium scindens,*^46^ but has been mapped to these taxa in our cohort. The BA conversion ability of the *Eggerthella lenta* was previously demonstrated *in vitro,* primarily focusing on the 3-, 4-, and 12-α HSDH genes.^18^ To our knowledge, *Eggerthella lenta* has not previously been reported to carry the *BaiB* gene and was considered to play only a minor role in secondary BA metabolism *in vivo.*^47^ However, in our study population, *Eggerthella lenta* is highly enriched in ALD patients and shows a high prevalence of MAGs containing the *baiB* gene, highlighting its potential contribution to the altered BA profile observed in ALD.

We identified 1,037 species as carriers of the 3-dehydro-bile acid Δ4,6-reductase *baiN* gene (Figure 5A). The *baiN* gene was experimentally characterized and identified in *Clostridium scindens*^48,49^, and homologs have been detected across the families *Ruminococcaceae*, *Lachnospiraceae*, and *Peptostreptococcaceae.*^50^ *Fusocatenibacter saccharivorans*, a member of *Lachnospiraceae,* showed a high prevalence in MAGs containing *baiN* but was depleted in ALD patients in our analysis and previous reports on liver cirrhosis.^51^

### Mediterraneibacter torques and Bacteroides thetaiotaomicron drive BA oxidation and epimerization in ALD

MAGs containing the BA oxidation and epimerization pathway-associated 7-α-hydroxysteroid dehydrogenase KEGG ortholog (*hdhA*, *hsdh*, K00076) were identified in 197 species, many of which also possess other key genes (Figure 5A). Numerous species carrying *hdhA* are enriched in ALD, with *Mediterraneibacter torques* and *Bacteroides thetaiotaomicron* showing a near-universal presence of *hdhA* across their MAGs and notable prevalence in ALD samples, underscoring their role in BA metabolism in ALD patients (Figure 5D). Previously, *hdhA* activity has been observed in *Clostridium*, *Eubacterium,* and *Ruminococcus* species.^52,53^ In our cohort, we found *hdhA* in several *Clostridium* species and *Blautia_A wexlerae.* However, although *hdhA* is ubiquitously present in various *Clostridium* species, its low prevalence in the GALAXY cohort suggests limited relevance to BA metabolism in ALD.

## Discussion

In this study, we observed increased bile acid (BA) abundance in the plasma of patients with alcohol-related liver disease (ALD), accompanied by downregulation of proteins involved in BA production and transport. Elevated BA concentrations were also evident in fecal samples, reflecting excessive BA excretion. We further identified that microbes alter secondary BA metabolism, leading to higher concentrations of TUDCA, GUDCA, and DCA. By mapping homologous genes, we pinpointed the microbial taxa most responsible for those changes in ALD, highlighting new targets for mechanistic studies.

### Downregulation of classical bile acid synthesis in ALD

Figure 6 presents an integrated view of host and microbial BA metabolism in ALD. Primary BA production of cholic acid (CA) and chenodeoxycholic acid (CDCA) from cholesterol is reduced in the liver, as demonstrated by decreased levels of biosynthetic enzymes in the classical pathway. Suppression of CYP8B1 has been shown to reduce CA synthesis and to increase the hydrophobicity of the BA pool via accumulation of 7α-hydroxy-4-cholesten-3-one (7αC4)^54^, an extremely hydrophobic and cytotoxic BA reported to cause liver cell damage.^55^ Deficiency in AKR1D1 leads to accumulation of toxic intermediates such as 3-oxo-Δ4 BAs and allo-BAs in the liver, which is associated with progressive intrahepatic cholestasis, hepatocyte injury, and apoptosis.^56^ To mitigate the excess of hepatotoxic BAs, compensatory mechanisms reduce BA accumulation in hepatocytes.^2^ For example, primary BA synthesis is regulated by the nuclear farnesoid X receptor (FXR) in the gut epithelium via a negative feedback mechanism.^12^ Gut microbes can inhibit BA production by producing FXR antagonists that regulate CYP8B1 expression, the rate-limiting enzyme in BA synthesis^13^. Excessive alcohol consumption in ALD disrupts the enzymatic detoxification, alters microbial composition,^57^ and compromises intestinal barrier function.^58,59^ This interplay between disrupted liver BA metabolism and gut microbial alterations, combined with impaired FXR signaling, likely exacerbates leaky gut and microbial translocation, creating a vicious cycle of liver injury and inflammation.

**Figure 6:**
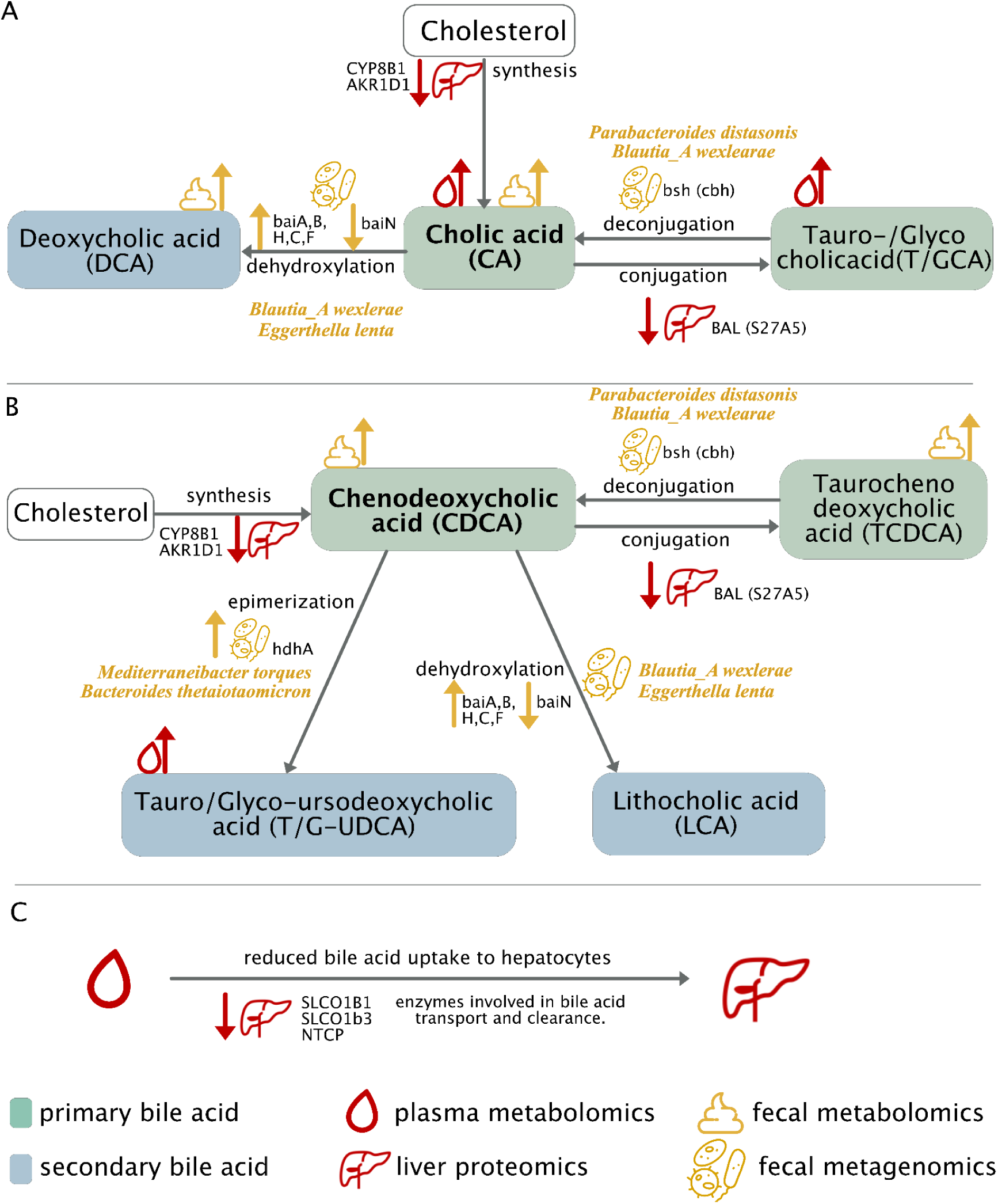
Combined representation of bile acid synthesis changes upon alcohol-related liver disease (ALD). **A-B**. The primary BAs cholic acid (A) and chenodeoxycholic acid (B) are metabolised by the secondary BA metabolism. **C.** Primary BA metabolism and transport by liver proteins. **A-C.** *Bsh*=bile salt hydrolase (also *cbh*=choloylglycine hydrolase), *hdhA*=7-alpha-hydroxysteroid dehydrogenase, *bai=*bai operon.

### Bile acid transport and the mechanism against hepatotoxic bile acids are disrupted

Our study shows dysregulation of several enzymes that are crucial for the BA transport across different compartments in ALD (Figure 6C). Downregulation of both SLCO1B1 and SLCO1B3, responsible for the hepatic uptake of BAs from blood, has been previously associated with Rotor syndrome, a condition characterized by high concentrations of plasma unconjugated BAs and bilirubin. These transporters compensate for the dysfunction of the main uptake transporter, the NA+-dependent taurocholate cotransporting polypeptide (NTCP).^60^ NTCP is recognized as the main uptake transporter for conjugated BAs from plasma to the liver, and several studies have demonstrated that its deficiency is related to highly elevated concentrations of conjugated BAs in the blood.^60–62^ In our study, NTCP was undetectable in more than 70% of the liver samples analyzed via MS-proteomics, as the levels were below the detection/quantification limit. The simultaneous downregulation of NTCP, SLCO1B1, and SLCO1B3 indicates impaired hepatic BA extraction, explaining the increased plasma concentrations of TCA, GCA, TUDCA, and GUDCA observed with fibrosis progression.

Loss of the bile salt export pump (BSEP) contributes to hepatocellular accumulation of toxic BAs, leading to intrahepatic cholestasis.^63^ However, the phosphatidylcholine transporter (ABCB4) was significantly upregulated with increasing liver fibrosis. ABCB4 facilitates the excretion of phospholipids into bile, forming mixed BA-phospholipid micelles that protect biliary epithelial cells from irritation due to high BA salt concentrations. The homeostasis of this protective synergy between BSEP and ABCB4 appears to be disrupted in alcohol-related fibrosis.^64^

### Contrasting changes in microbial bile acid dehydroxylation genes reflect dysbiosis and community-level metabolic dysfunction

Alcohol overuse was shown to induce translocation of microbes and their metabolites, including BAs, due to compromised intestinal barrier function.^58,59^ The most cytotoxic secondary BAs are LCA and DCA; the latter was found enriched in the feces of ALD patients, possibly contributing further to intestinal permeability.^65^ These BAs have been implicated in cell membrane disruption, bile duct damage, inflammation, and cholestasis, causing hepatocyte injury, liver fibrosis, and liver cancer.^66–68^ The *bai* operon governs dehydroxylation of primary BAs to LCA and DCA in the gut, and is well described, for example, in *Clostridium* species^49^. However, *Clostridium* species were not a dominant carrier of *bai* genes in our cohort. Instead, *Eggerthella lenta* and *Blautia_A wexlerae* seem to assume key roles in BA dehydroxylation in ALD but lack the full enzymatic pathway. We found early *bai* genes to be enriched, whereas the late-pathway gene *baiN*, although highly abundant overall, strongly decreased with disease progression, indicating a metabolic bottleneck. This community-level dysfunction likely results from microbial dysbiosis induced by alcohol overuse. We hypothesize that impaired conversion to secondary BAs and accumulation of toxic intermediates may exacerbate intestinal barrier disruption, fostering a vicious cycle of increased permeability, bacterial products, and bile acid translocation to the liver, and hepatic inflammation.^69^

### In the early fibrosis stage, epimerization of bile acids detoxifies the bile acid pool

The secondary BAs UDCA and TUDCA exhibit low toxicity and anti-inflammatory properties and are clinically used to treat cholestatic liver disease and biliary cirrhosis.^70,71^ We observed increased TUDCA levels in blood and elevated abundance of the *hdhA* gene, responsible for UDCA and TUDCA production, peaking at fibrosis stage two. However, *hdhA* levels declined in cirrhosis and advanced fibrosis across cohorts, and TUDCA was further depleted in advanced fibrosis in validation cohort 1. This U-shaped pattern, also noted for other genes, proteins, and metabolites in ongoing studies within the consortium, suggests an initial compensatory detoxification response that diminishes at later disease stages, potentially due to microbial dysbiosis and increased intestinal permeability. In summary, the increased pool of hydrophobic BA in the early fibrosis appears to trigger this detoxification pathway, which we hypothesize is lost in advanced fibrosis as a consequence of microbial dysbiosis and intestinal barrier dysfunction associated with ALD.

We also found that azelaic acid levels in feces decreased with fibrosis progression. Azelaic acid is known to have anti-inflammatory, antioxidant, and antibacterial properties. Further, azelaic acid has demonstrated anti-atherosclerotic action,^72^ and protects the liver against oxidative stress induced by ethanol and high-fat diets.^73^

Our cross-sectional study design presents a limitation to inferring cause and effect in the findings, and thus, our findings are to be interpreted as associations. Moreover, the small sample size in the validation cohorts sets a limit on the power of validation. In the discovery cohort, BA levels across fibrosis stages were compared to levels from healthy controls, while in the validation cohorts, early and late fibrosis stages were compared. This difference in comparison approaches may account for some discrepancies, such as the observed reduction of TCDCA at stage four. In addition, the microbial functional analyses rely on gene ontology databases, which often lack mechanistic proof for individual taxa and are limited to genes available in the databases. The strengths of the present study, however, lie in the detailed characterization of the liver disease and deep phenotyping with various omics technologies. Unlike prior studies focusing on individual aspects of BA metabolism, we present a comprehensive integrated analysis of the whole gut-liver-BA axis and its disruption in ALD.

## Abbreviations

ALD: Alcohol-related liver disease
BAL: Bile acid CoA ligase
*bai*: Bile acid-induced
BMI: Body mass index
BSEP: Bile salt export pump
bsh: Bile salt hydrolase
CA: Cholic Acid
CDCA: Chenodeoxycholic acid
DCA: Deoxycholic acid
GCA: Glycocholic acid
GTDB: Genome taxonomy database
GUDCA: Glycoursodeoxycholic acid
FXR: Farnesoid X receptor
HC: Healthy control
HdhA: 7-alpha-hydroxysteroid dehydrogenase
KEGG: Kyoto encyclopedia of genes and genomes
LCA: Lithocholic acid
MAG: Metagenome-assembled genome
MELD: Model of end-stage liver disease
MetaG: Metagenomics MetaT Metatranscriptomics
mOTU: Metagenomic operational taxonomic units
NTCP: NA+ dependent taurocholate cotransporting polypeptide 28
TCA: Taurocholic acid
TCDCA: Taurochenodeoxycholic acid
TUDCA: Tauroursodeoxycholic acid
UHPLC-MS/MS: Ultra-high performance liquid chromatography-tandem mass spectrometry

## Acknowledgments

We thank all members of the GALAXY and MicrobLiver consortia for fruitful discussions and support. We would like to express our sincere gratitude to Lise Ryborg, Louise Skovborg Just, and Ida Falk Villesen for their invaluable contributions in managing the administrative aspects of the consortia. This work was supported by funding from the European Union’s Horizon 2020 research and innovation program under grant agreement number 668031 (GALAXY). This work reflects only the authors’ view, and the European Commission is not responsible for any use that may be made of the information it contains. The study was also supported by the Novo Nordisk Foundation through a Challenge Grant “MicrobLiver” (grant number NNF15OC0016692), by the EMBL International PhD Programme (M.I.K.), by the Deutsche Forschungsgemeinschaft (DFG, German Research Foundation) - project number 460129525 (NFDI4Microbiota), and by EMBL IT Services HPC resources. Finally, we would like to express our deepest gratitude to all participants in the cohorts presented.

## Author contributions

Conceptualization: M.I.K., A.Z., M.T., C.L.Q., P.B., A.K., T.H.; data curation: A.Z., T.S., L.N., A.N.F., W.A., C.S., D.P., C.Y.K., A.W., M.K., M.I.K., H.B.J., S.N.; data analysis: A.Z., T.S., M.I.K., S.N.; sample collection: M.T., A.K., J.K.H., M.I.; sample processing A.Z., D.H., L.N., M.I.K.; writing: A.Z., M.I.K., C.L.Q., M.K., D.P.; visualization: M.I.K., A.Z., T.S., K.S.; supervision: C.L.Q., K.S., M.K., P.B.; funding acquisition: A.K., T.H., C.L.Q., P.B.

All authors discussed the results, reviewed the manuscript, and approved the final manuscript.

## Data availability

Shotgun metagenomic data sequenced in the GALAXY/MicrobLiver consortia are publicly available in the European Nucleotide Archive under the accession numbers of BioProject: PRJEB76661 (GALA-ALD), BioProject: PRJEB76664 (GALA-HP), PRJEB76667 (Validation 1, GALA-RIF), and PRJEB76668 (Validation 2, GALA-POSTBIO). Clinical contextual data, proteomics, and targeted metabolomics data cannot be made publicly available due to the higher need to maintain patient confidentiality. Averaged protein levels in the liver proteomes have been deposited in the GitHub repository (https://github.com/llniu/ALD-study, subfolder ALD-App). Permission to access and analyze data can be obtained following approval from the Danish Data Protection Agency and the ethics committee for the Region of Southern Denmark. The study protocol, standard operating procedures, and patient information are also available upon request.

## Clinical trial number

GALAXY discovery cohort: Danish Data Protection Agency nos. 13/8204, 16/3492 and 18/22692; and Odense Patient Data Exploratory Network under study identification nos. OP_040 and OP_239

Validation cohort 1: EudraCT number 20214-001856-51

Validation cohort 2: ClinicalTrial.gov ID NCT03863730

## Supplementary Material

**Supplementary Table 1:**
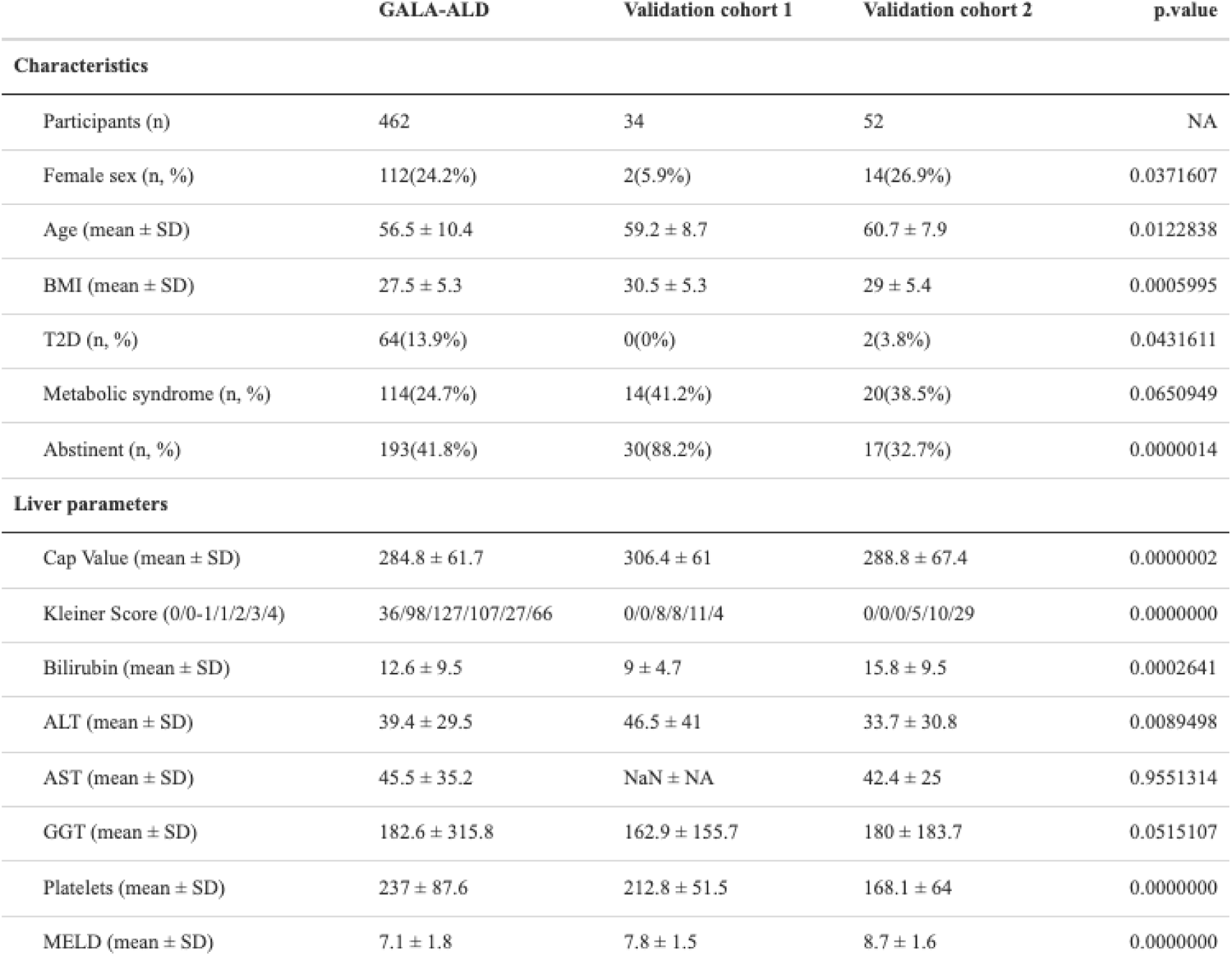
Characteristics of the GALAXY alcohol-related liver disease (ALD) cohort in comparison to the Validation 1 and 2 cohorts. T2D = Type 2 Diabetes, Cap = Controlled attenuation parameter, ALT = alanine transaminase, AST = aspartate aminotransferase, GGT = gamma-glutamyl transferase, MELD = model of end-stage liver disease. P-values report the significance of the differences between the ALD and HC cohorts tested by the Kruskal-Wallis test for continuous variables and the chi-squared test for categorical variables.

|  | GALA-ALD | Validation cohort 1 | Validation cohort 2 | p.value |
| --- | --- | --- | --- | --- |
| **Characteristics** |  |  |  |  |
| Participants (n) | 462 | 34 | 52 | NA |
| Female sex (n, %) | 112(24.2%) | 2(5.9%) | 14(26.9%) | 0.0371607 |
| Age (mean ± SD) | 56.5 ± 10.4 | 59.2 ± 8.7 | 60.7 ± 7.9 | 0.0122838 |
| BMI (mean ± SD) | 27.5 ± 5.3 | 30.5 ± 5.3 | 29 ± 5.4 | 0.0005995 |
| T2D (n, %) | 64(13.9%) | 0(0%) | 2(3.8%) | 0.0431611 |
| Metabolic syndrome (n, %) | 114(24.7%) | 14(41.2%) | 20(38.5%) | 0.0650949 |
| Abstinent (n, %) | 193(41.8%) | 30(88.2%) | 17(32.7%) | 0.0000014 |
| **Liver parameters** |  |  |  |  |
| Cap Value (mean ± SD) | 284.8 ± 61.7 | 306.4 ± 61 | 288.8 ± 67.4 | 0.0000002 |
| Kleiner Score (0/0-1/1/2/3/4) | 36/98/127/107/27/66 | 0/0/8/8/11/4 | 0/0/0/5/10/29 | 0.0000000 |
| Bilirubin (mean ± SD) | 12.6 ± 9.5 | 9 ± 4.7 | 15.8 ± 9.5 | 0.0002641 |
| ALT (mean ± SD) | 39.4 ± 29.5 | 46.5 ± 41 | 33.7 ± 30.8 | 0.0089498 |
| AST (mean ± SD) | 45.5 ± 35.2 | NaN ± NA | 42.4 ± 25 | 0.9551314 |
| GGT (mean ± SD) | 182.6 ± 315.8 | 162.9 ± 155.7 | 180 ± 183.7 | 0.0515107 |
| Platelets (mean ± SD) | 237 ± 87.6 | 212.8 ± 51.5 | 168.1 ± 64 | 0.0000000 |
| MELD (mean ± SD) | 7.1 ± 1.8 | 7.8 ± 1.5 | 8.7 ± 1.6 | 0.0000000 |

**Supplementary Figure 1:**
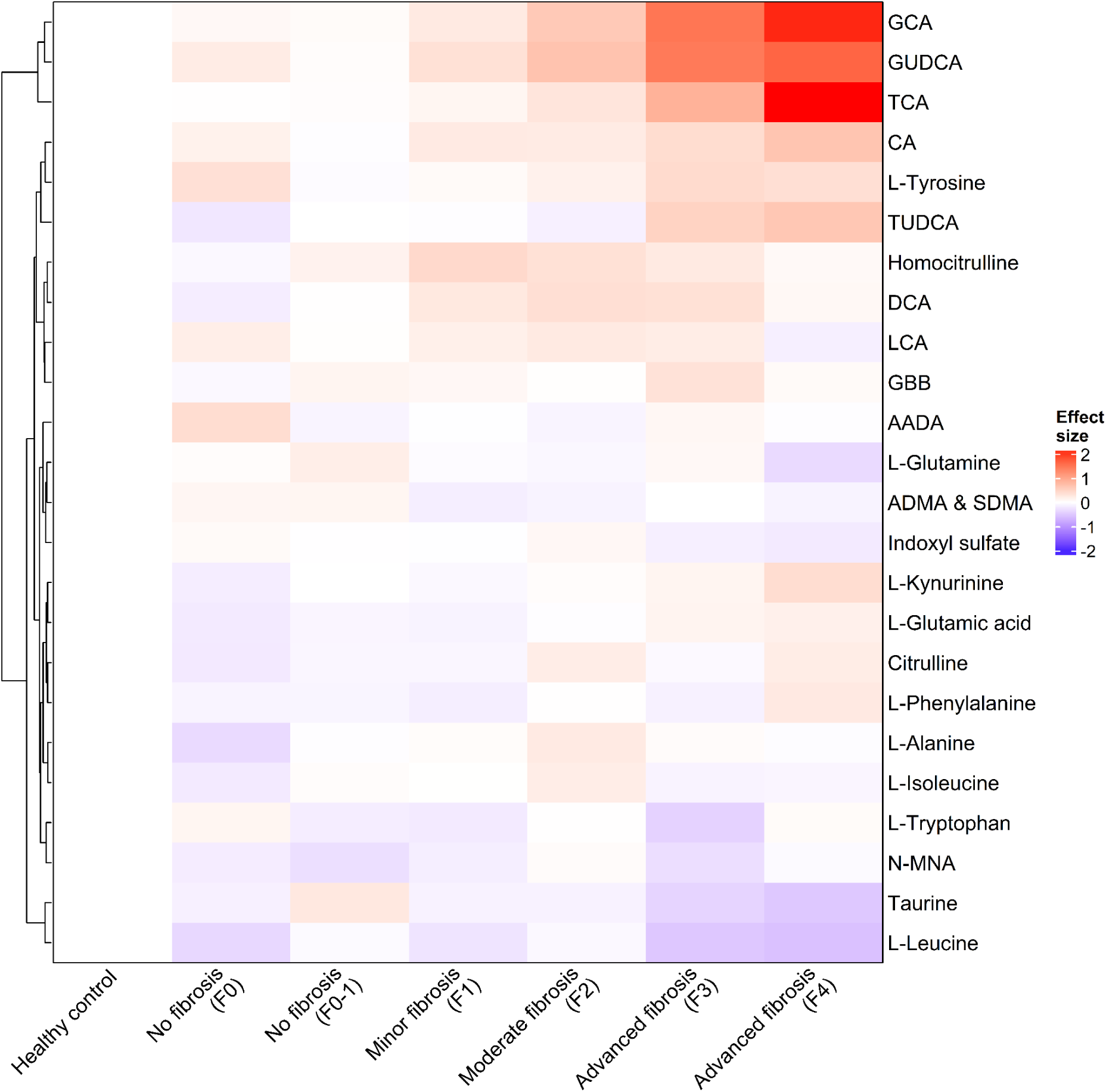
Blood metabolite levels according to fibrosis stage. Heatmap of the standardized effect size of metabolite level as compared to healthy controls with all measured metabolites in blood. Metabolites are shown in rows (ordered by hierarchical clustering), fibrosis stages in columns, and the respective effect size in color (red: increase; blue: decrease).

**Supplementary Figure 2:**
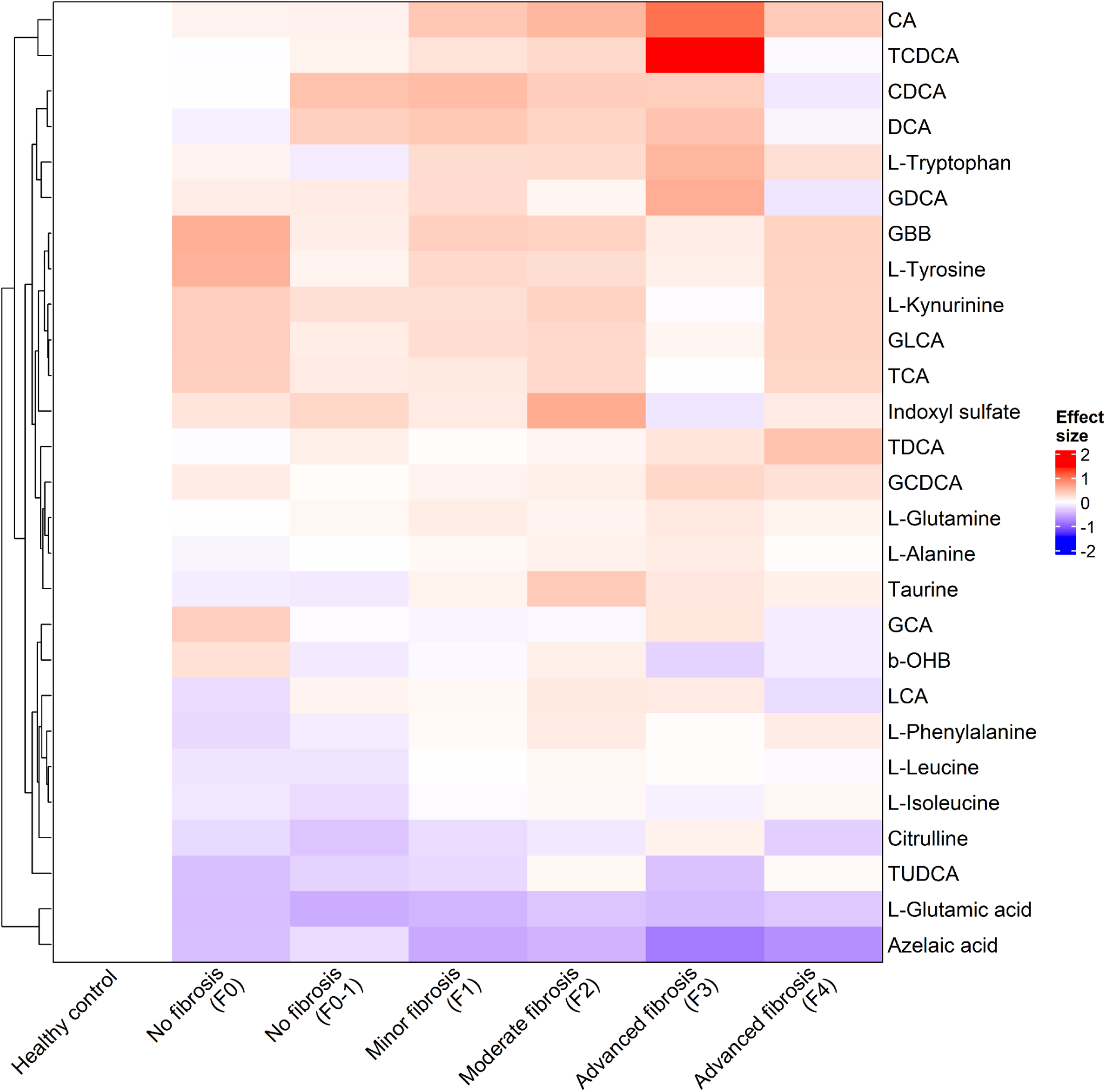
Fecal metabolite levels according to fibrosis stage. Heatmap of the standardized effect size of metabolite level as compared to healthy controls with all measured metabolites in feces. Metabolites are shown in rows (ordered by hierarchical clustering), fibrosis stages in columns, and the respective effect size in color (red: increase; blue: decrease).

**Supplementary Figure 3:**
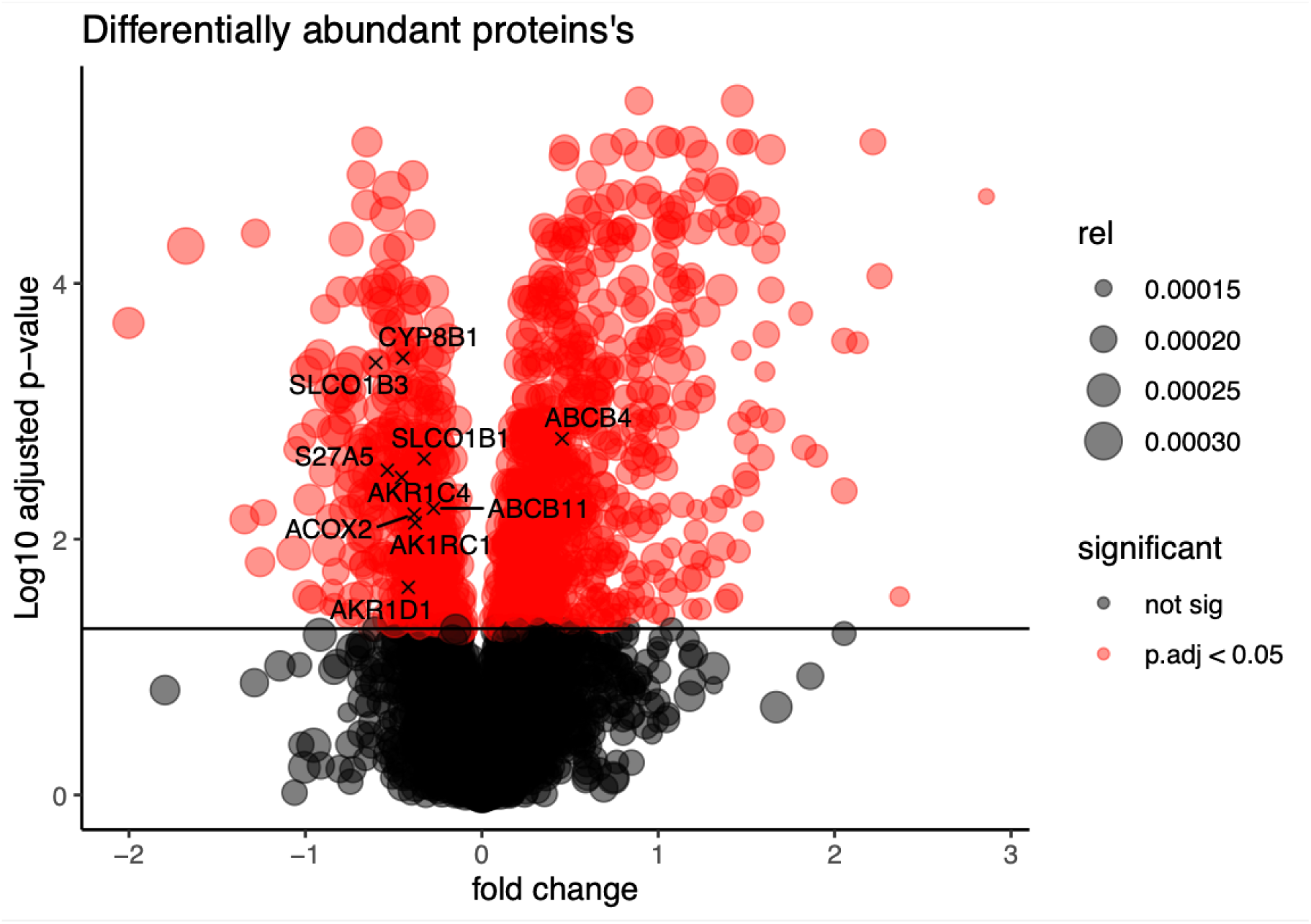
Differential abundance analysis of the liver proteomics dataset comparing early fibrosis (F0-1) with late fibrosis (F2-4). Proteins in red are significantly expressed after correcting for false positives using the Benjamini-Hochberg correction. Points with labels are related to BA synthesis and transport.

**Supplementary Figure 4:**
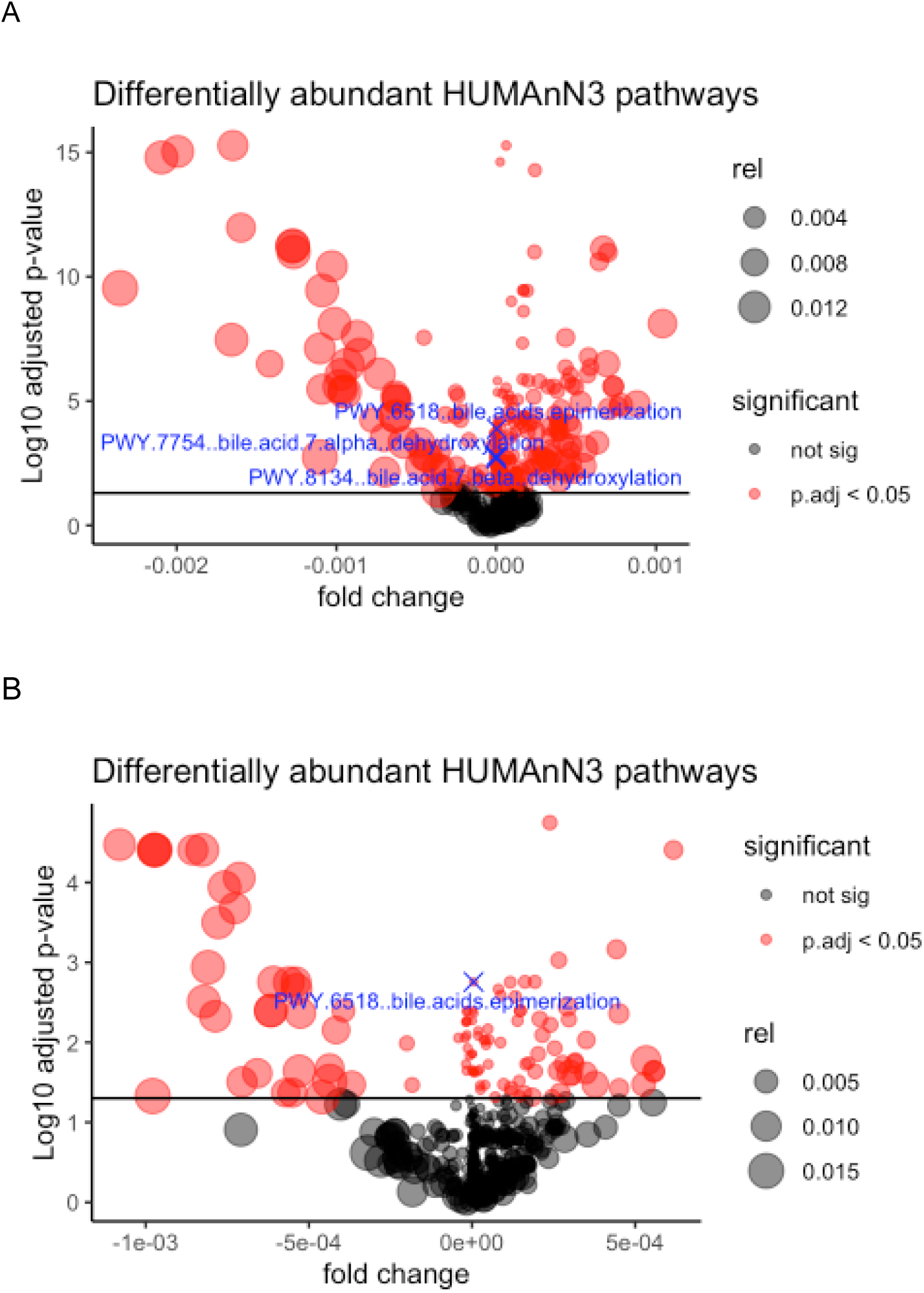
Differential abundance analysis of the microbial pathways derived from metagenomic sequencing of fecal samples. The test compares the ALD with the HC samples (A) and within the ALD cohort, the early fibrosis (F0-1) with late fibrosis (F2-4) (B). Points in red are significantly changing pathways between the comparisons after adjusting for false positives using the Benjamini-Hochberg correction. Points with labels secondary BA metabolism pathways.

**Supplementary Figure 5:**
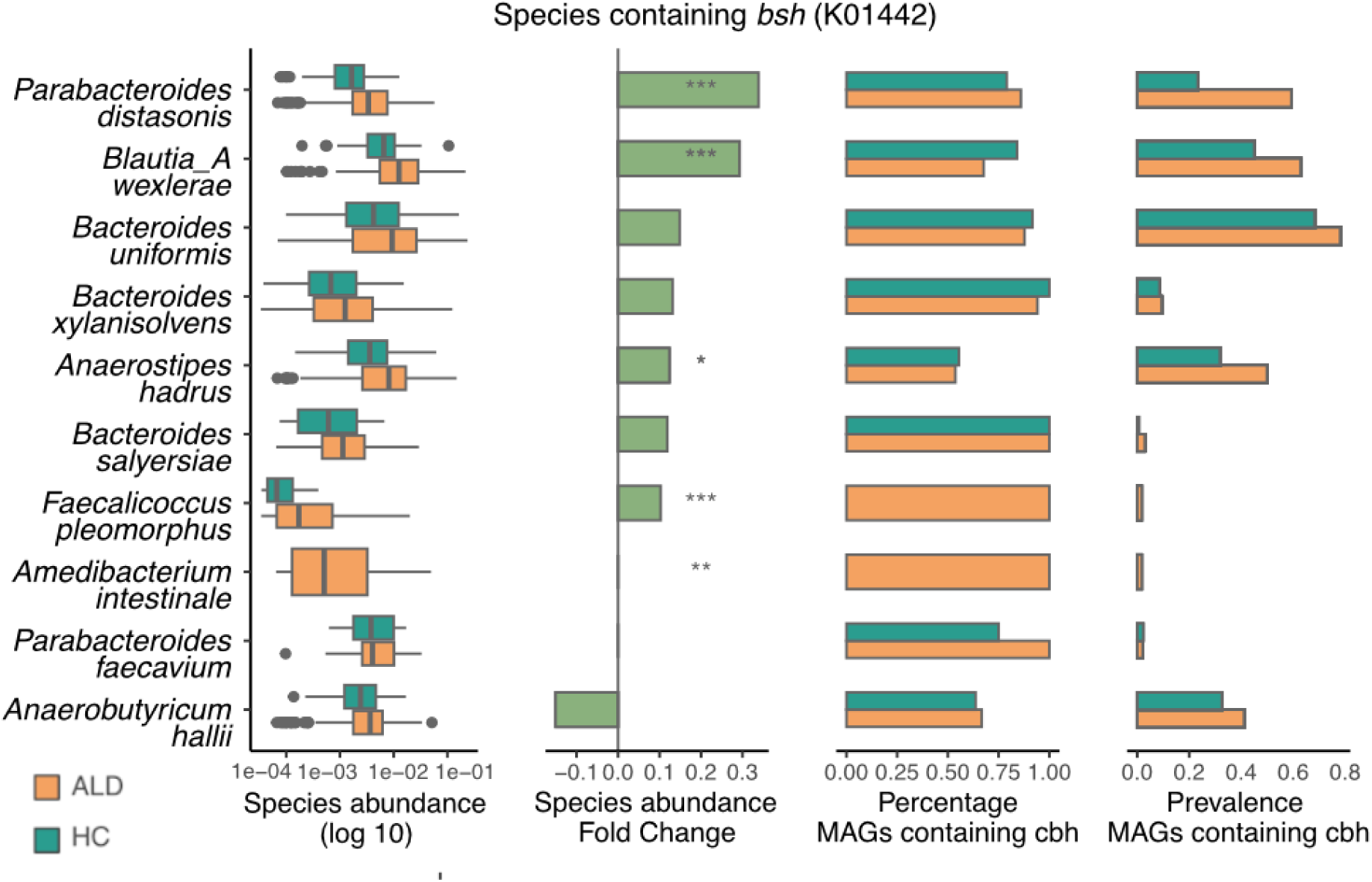
Microbial taxa for the secondary bile acid metabolism deconjugation pathway facilitated by *bsh.* The species abundance, species fold change between healthy and ALD samples, the gene-carrying frequency of MAGs, and the prevalence of gene-carrying MAGs in the cohort collectively estimate the importance of the taxa in ALD.

